# Age and the relation of common neuropathologies to dementia in Brazilian adults

**DOI:** 10.64898/2026.02.11.26346038

**Authors:** Jose M. Farfel, Sukriti Nag, Ana W. Capuano, Maria Carolina M. Sampaio, Victoria N. Poole, Robert S. Wilson, David A. Bennett

## Abstract

**Background:** Community-based clinical-pathologic studies have been instrumental to examine the association of Alzheimer’s disease and related disorders (AD/ADRD) with age and dementia in very-old non-Latino Whites. Here, we show the age distribution of four AD and three additional common neuropathologies across the adult lifespan and examine their relation to dementia and cognitive impairment in old and young Brazilian adults.

**Methods:** We examined 5,376 brains from decedents age 18 years or older (52.5% male, 39.8% Black), from the Pathology, Alzheimer’s and Related Dementias Study (PARDoS), collected between July 2021 and September 2025. Clinical diagnoses were rendered by a clinician who reviewed the Informant Questionnaire on Cognitive Decline in the Elderly (IQCODE), informant-based Clinical Dementia Rating (CDR) Scale, and other selected data. Four indices of AD including β-amyloid deposits (Thal stage), PHF-tau tangles (Braak stage), neocortical phosphorylated plaques and AD neuropathologic change (ADNC), and three other common neuropathologies, i.e., Lewy-body disease (LBD), chronic gross infarcts, and cerebral amyloid angiopathy (CAA) were assessed. Logistic regression was used for associations of pathologies with clinical diagnoses, adjusting for demographics.

**Results:** Intermediate to high ADNC were first found as early as the fourth decade. Chronic gross infarcts were found in one-fifth of the brains of young adults. Intermediate to high ADNC, limbic and neocortical LBD, chronic gross infarct and moderate to severe CAA were associated with dementia and cognitive impairment (CI) in older adults with mixed pathologies being the most common. Intermediate to high ADNC was associated with CI but not dementia in young adults, whereas, chronic gross infarcts were associated with both CI and dementia in young adults; overall, mixed pathologies were a small minority.

**Conclusion:** In a community-based, clinical-pathologic study including 5300+ brains from diverse Brazilians, we show that AD and other common pathologies frequently begin in young adulthood. In older adults, mixed pathologies are most commonly associated with dementia, whereas in young adults a single pathology, most commonly chronic gross infarcts rather than ADNC is related to dementia.

## Introduction

Clinical-pathologic studies have been instrumental for brain aging and dementia research by characterizing in a more detailed and specific manner Alzheimer’s disease (AD) and related disorders (ADRD) [1–3] and showing that AD pathology most frequently develops together with one or more co-morbid neuropathologies. [4–8] Clinical-pathologic studies commonly enroll convenience samples composed of patients referred to dementia clinics with severe or atypical clinical presentations. [9–12] Community-based studies are relatively few [13–22] and recruit mostly very-old, highly educated, non-Latino white women autopsied after the age of 85. As a result, only a few studies include the distribution of ADRD across decades [23–25], but with very limited sample size of age < 80. One exception is a different large clinical-pathologic study also conducted in Brazil with a mean age at death of 74. [26] However, we are unaware of any community-based studies that examined the relation of common neuropathologies to clinical diagnoses among those age 18-64. Thus, there remains a need for clinical-pathologic studies in populations that represent greater diversity, especially a broader age range.

The Pathology, Alzheimeŕs and Related Dementia Study (PARDoS) is an ongoing community-based, clinical-pathologic study enrolling Brazilian decedents age 18 years or older, with a broad education and socioeconomic range, a relatively balanced sex and race distribution, who died from non-forensic causes of death and received an autopsy in one of two Autopsy Services or a tertiary care Hospital in the State of Sao Paulo starting in July 31, 2021. [27] PARDoS fills a major demographic gap in the aging and dementia clinical-pathology portfolio of studies. Here, we present the age-distribution of dementia and mild cognitive impairment (MCI), four AD indices and three-other common neuropathologies across more than eight decades in 5300+ decedents, including a large number under age 65. We show the distribution of single or mixed neuropathologies in dementia, MCI and no cognitive impairment (NCI), and examine the association of AD/ADRD with dementia and cognitive impairment in both young and older adults.

## Methods

### Study participants

PARDoS is based at the Hospital do Servidor Publico Estadual (HSPE) of IAMSPE, a healthcare network for the public servants of the State of Sao Paulo and their families. [27] PARDoS recruits decedents aged 18 years or older with a legal representative providing consent for the study. The study prioritizes, proxy-reported Black/Mixed race and White race with 8 or fewer years of education. Decedents born in Paraná, Santa Catarina and Rio Grande do Sul, the three southern states of Brazil where African ancestry is very limited are low priority. Forensic cases, cases retained by the pathologist for further investigation, and cases with advanced autolysis are ineligible.

A two-step ethical approval is required in Brazil for projects funded by foreign institutions. IAMSPE local ethics committee (CEP-IAMSPE) approved PARDoS as a new study in May, 2020 and the Comissão Nacional de Ética em Pesquisa (CONEP), the Brazilian national ethics committee approved the study in September, 2020. Informants of decedents are considered research subjects in Brazil and all provided written informed consent.

Participants were recruited from three sites. The Santo Andre Autopsy Service (Santo Andre SVO), located southeast and the Guarulhos Autopsy Service (Guarulhos SVO), located northeast of the city of Sao Paulo, and HSPE. Due to the COVID pandemic brain collection started on July, 2021 at Santo Andre, August, 2022 at Guarulhos, and August, 2023 at HSPE after obtaining approval by the public health authorities for a newly built autopsy suite.

From July 31, 2021 to September 29, 2025, PARDoS nurses at the three sites obtained consent from 6,977 legal representatives. Of these, 10.4% of decedents were ineligible, mostly due to retention of the brain by the autopsy service pathologist. PARDoS collected brains from 6,252 decedents in this period. Supplementary figure 1 shows when and from which site these cases were collected. Through September 29, 2025, data for classifying at least one of the neuropathologies examined in the manuscript were available in 5,376. Their mean age at death was 72.0 years (SD: 14.4 years, range 18 to 108), 30.4% were under age 65, 52.5% were male, 39.8% were Black/Mixed, 57.7% were Whites and 1.8% other races, primarily Asian (mostly Japanese). Their mean education was 6.5 years (SD: 4.4 years). Table 1 summarizes the sample characteristics in 18-64 and 65+ decedents.

**Table 1.** Sample characteristics in younger and older adults.

| CHARACTERISTIC | N | 65+ Decedents | 18-64 Decedents | Total |
| --- | --- | --- | --- | --- |
| Age at death, mean (SD) | 5376 | 79.6 (8.7) | 54.6 (8.4) | 72.0 (14.4) |
| Male sex, n (%) | 5376 | 1771 (47.3) | 1052 (64.3) | 2823 (52.5) |
| Black race, n (%) | 5373 | 1348 (36.1) | 792 (48.4) | 2140 (39.8) |
| Education, mean (SD) | 5373 | 5.7 (4.3) | 8.3 (4.1) | 6.5 (4.4) |
| Thal Staging $\geq 3$, n (%) | 5338 | 1620 (43.6) | 145 (8.9) | 1765 (33.1) |
| Braak Staging $\geq 3$, n (%) | 5343 | 2657 (71.5) | 222 (13.6) | 2879 (53.9) |
| Moderate to Severe Phosphorylated Plaques, n (%) | 5235 | 688 (18.7) | 19 (1.2) | 707 (13.5) |
| Intermediate to High ADNC, n (%) | 5338 | 1506 (40.5) | 57 (3.5) | 1563 (29.3) |
| Limbic and Neocortical LBD, n (%) | 5296 | 570 (15.5) | 56 (3.5) | 626 (11.8) |
| Gross Infarcts, n (%) | 5376 | 1216 (32.5) | 367 (22.4) | 1583 (29.4) |
| Moderate to Severe CAA, n (%) | 5358 | 1006 (27.0) | 84 (5.2) | 1090 (20.3) |

### Clinical diagnoses

An interview with informants to obtain ante-mortem clinical information about the decedents was conducted by PARDoS nurses in 88.9% and the clinical diagnoses were completed in 79.8% of the brains collected with interviews and diagnostic classification ongoing. The 26-item Informant Questionnaire of Cognitive Decline in the Elderly (IQCODE) is used to determine cognitive decline in the years prior to death [28] and the Clinical Dementia Rating Scale (CDR) is used as a measure of cognitive impairment. [29] The IQCODE and CDR were combined by an algorithm and an experienced clinician confirmed or modified the algorithm following review of relevant information in the nurses’ informant interview notes and other data from the interview, e.g., stroke. The final diagnoses were no cognitive impairment (NCI), mild cognitive impairment (MCI), dementia, and cognitive impairment no dementia (CIND) for persons with lifelong cognitive impairment but no cognitive decline. Cognitive impairment combines decedents diagnosed with dementia or MCI. CIND cases, present primarily in young decedents, were excluded in these analyses. More information about the interview and diagnostic process was previously reported. [27] Through September 29, 2025, data for clinical classification was available in 4818 (89.6%) of the cases on whom the brain was obtained.

### Demographic data

Proxy-declared sex and race classified as Black, Mixed, White, Asian, and Native-American were obtained. Blacks and the Mixed were merged in one category. Education was reported by the informants as the number of years of formal school attendance.

## Neuropathologic examination

### Post-Mortem Procedures

Experienced pathology technicians performed post-mortem brain procurement and pathology data collection with quality control conducted by two neuropathologists. The procedures are similar to those used for the Religious Orders Study and Rush Memory and Aging Project (ROSMAP) at Rush Alzheimer’s Disease Center, modified to accommodate the large number of autopsies.[21] Procedures follow recent guidelines for the neuropathologic diagnosis of ADRD. [1–3,32] Thal β-amyloid staging used immunohistochemistry (IHC) (4G8, 1:9000 Covance Labs, Madison, WI) on neocortex (midfrontal and middle temporal cortices), mid-hippocampus, basal ganglia, midbrain and cerebellum. Using anti-PHFtau IHC (AT8, 1:2000; Thermoscientific, Waltham, MA), we quantitated PHF-tau neurofibrillary tangles (NFT) in the entorhinal cortex, mid-hippocampus and midfrontal, middle-temporal and calcarine cortices. The Braak score is derived from the NFT counts. Also using anti-PHFtau IHC, we quantitated phosphorylated plaques (PP) in midfrontal and middle-temporal cortices. For NFT and PP, the region with the greatest density of each finding was selected and the total number of NFT, and separately PP, were quantitated at a magnification of 100X in a one mm^2^ area. A semiquantitative scale was used to classify PP in none, mild, moderate and severe. The AD neuropathologic change (ADNC) assessment included the ABC score, derived from the Thal β-amyloid stage, the Braak score (derived from NFT counts) and the CERAD score (derived from the PP counts), respectively. The neuropathologic diagnosis of AD required intermediate to high ADNC following the NIA-AA criteria [1].

Cerebral amyloid angiopathy (CAA) was assessed with β-amyloid IHC using a previously established scale in meningeal and parenchymal vessels examined in midfrontal and calcarine cortices [33]. Lewy bodies (LB) were identified by alpha-synuclein IHC (pSyn, 1:20000; Wako Chemicals, Richmond, VA) in the olfactory bulb, midbrain, entorhinal cortex, amygdala and midfrontal and middle temporal cortices. The Lewy body stage is described olfactory only, amygdala predominant, nigral predominant, limbic type or neocortical type using modified staging criteria [34]. Gross infarcts were identified during brain dissection, and size, age and location were recorded and additional tissue blocks from each infarct collected for histologic confirmation [35].

Given the number of brains that PARDoS collects daily and the fact that many were young adults without pathology, several steps were implemented to streamline data generation and improve throughput. One step was to prioritize data collection for AD, LBD, gross infracts and CAA over other neurodegenerative and vascular pathologies which are ongoing. Here, we present data from these four pathologies. Data collection for TDP-43, hippocampus sclerosis, microinfarcts and arteriolosclerosis will be published at a later date.

### Statistical Analysis

Initial analysis includes careful examination of the distribution and correlation structure of the neuropathologies, along with clinical and demographic variables First, we show the age-distribution of dementia, MCI, four AD neuropathologic indices (β-amyloid deposits with Thal staging, NFT with Braak staging, semiquantitative scale for PP and AD neuropathologic change (ADNC) according to NIA-AA criteria), and three additional common neuropathologies (chronic gross infracts, LBD and CAA). We first describe the age-distribution of ADRDs using diagnostic criteria described in guidelines or previous publications (Thal stage ≥3, Braak stage ≥ 3, moderate to frequent PPs, intermediate to high ADNC, chronic gross infarcts, limbic and neocortical predominant LBD and moderate to severe CAA) [30–35]. Next, we examine when AD and other neuropathologies start in the brain, describing the age-distribution of early neuropathologic changes (Thal stage 1-2, Braak stage 1-2, sparse PPs, low ADNC, olfactory and nigra predominant LBD and mild CAA).

We present the clinical-pathologic analyses in the 65+ decedents consistent with other community-based studies enrolling older adults for comparison. The frequency of AD and the three additional neuropathologies occurring as a single pathology or in combination in participants with dementia, MCI and NCI is shown. Univariate analysis examined differences in neuropathologic variables in decedents with and without dementia using chi-square tests when appropriate for categorical variables and unpaired t tests for continuous variables. Next, we fit logistic regression models for (1) dementia versus no dementia and (2) cognitive impairment (dementia or MCI versus NCI), excluding CIND, with terms for the neuropathologies and adjusting for age at death (centered at its mean), sex, race and education (centered at its mean). First, we fit these models including the three AD indices deriving ADNC entered individually and then simultaneously into one model. Next, we fit these models including ADNC, chronic gross infarcts, limbic and neocortical predominant LBD and moderate to severe CAA, entered individually and then simultaneously into one model. We repeated the clinical-pathologic analyses for ages 18-64 decedents presenting novel pathologic data not explored in prior studies. A nominal p value < 0.05 determined statistical significance. The analyses were performed using SAS/STAT software, version 9.4 [SAS Institute Inc., Cary, NC].

## Results

### Age-distribution of pathologic ADRD

The distribution of β-amyloid deposits, NFT density and neocortical PPs are described in Supplementary Figure 2 and Supplemental material. The neuropathologic diagnosis of Intermediate to high ADNC was first observed about age 35 (figure 1a). It was less than 10% from the mid-fourth to the mid-seventh decades. It then increased rapidly to more than 10% in the seventh, a quarter in the eighth and more than half of the brains at the ninth and tenth decades.

**Figure 1.**
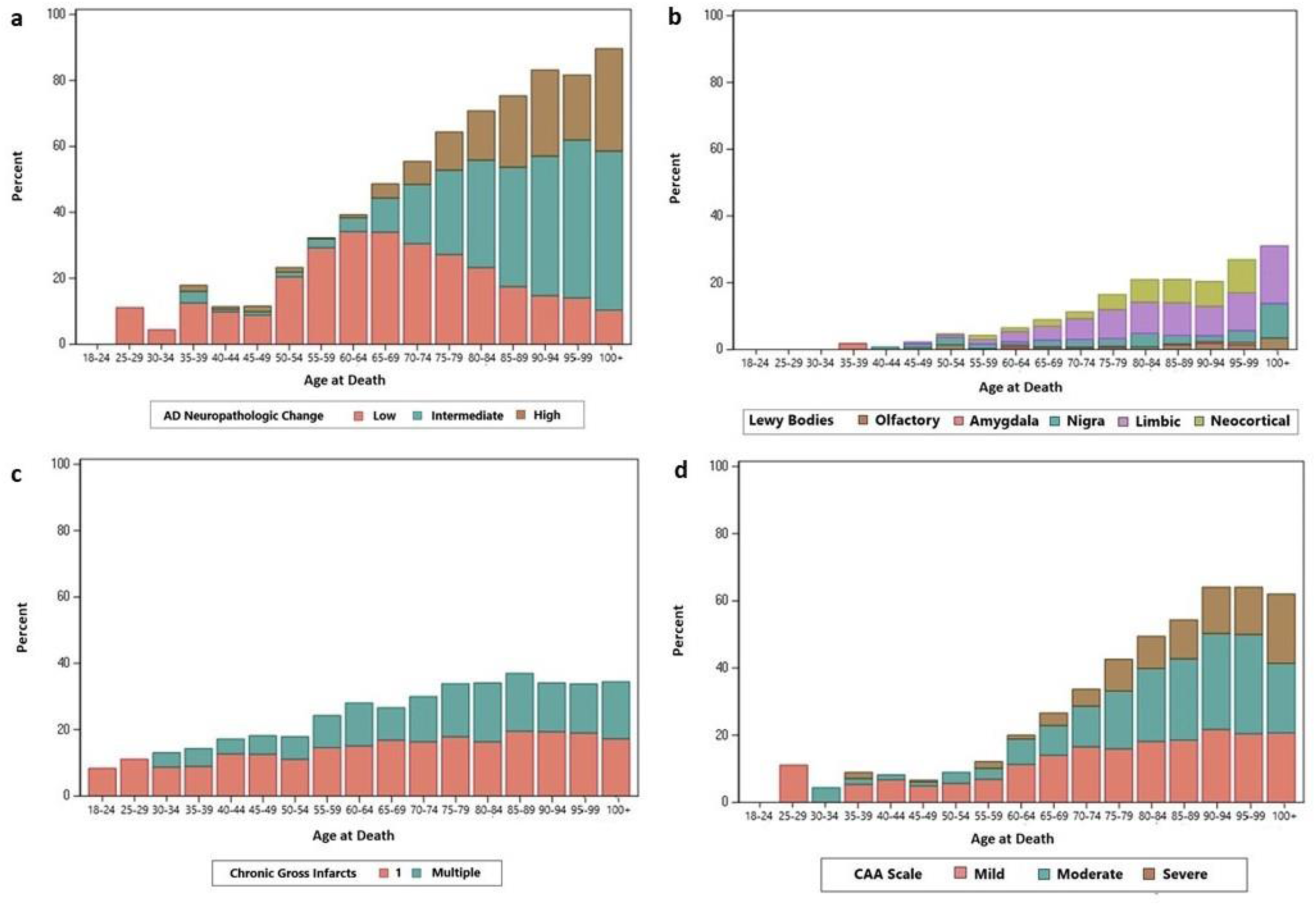
Age-distribution of ADNC and three other common neuropathologies.

The age-distribution of three non-AD neuropathologies, including LBD, chronic gross infarcts and CAA is shown in Figure 1b-d. Limbic and/or neocortical predominant LBD were first seen in the fifth decade, but very rare before the age of 65, present in over 5% in the mid seventh decade, one-tenth in the eighth and one-fifth in the ninth decade. Not surprisingly, chronic gross infarcts were found in the brains from the youngest decedents (ages 18-25) in this study of diverse Brazilians who had a natural cause of death. It was present in about 10% of the brains in the third and fourth decade, then increased rapidly to one-fifth in the fifth and sixth decades and a quarter to nearly 40% of the brains afterwards. Moderate-severe CAA was first seen in the third decade, although it remained rare between the fourth and mid-sixth decade, it then increased rapidly to 10% in the seventh, a quarter in the eighth and two-fifths in the ninth decade.

We show the upset plots for no, single, and mixed pathologies separately for age 65+ and age 18-64 (supplementary figures 3a and 3b). Among those age 65+, 36% had one pathology and 33% had two or more pathologies. The remaining 31% had no pathologic diagnosis. Among those age 18-64, 25% had one pathology and less than 5% had two or more pathologies. The remaining 70% had no pathologic diagnosis.

### Age-distribution of dementia and MCI

The age-distribution of dementia and cognitive impairment in the subset for whom diagnostic classification was complete is shown in Figure 2. Dementia was not found younger than the fourth decade, was rare in the fourth and fifth decades when it varied from few percent to more than 5%. It was present in one-tenth in the sixth and seventh decade, a quarter in the eight and two fifths in the ninth decade. MCI also was not present before the fourth decade, rare in the fourth decade, over 5% in the fifth and mid-sixth decades and one-tenth afterwards.

**Figure 2.**
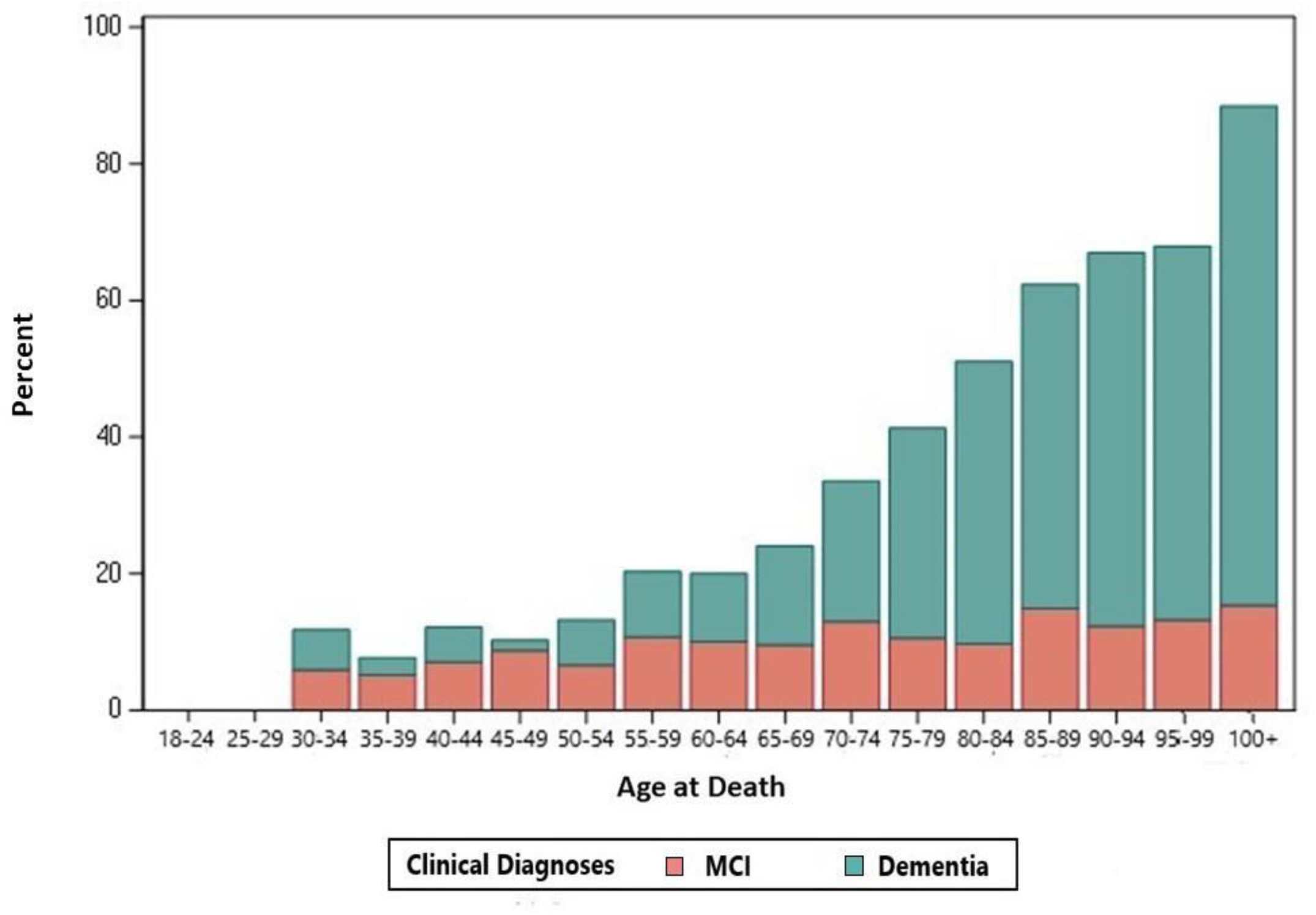
Age-distribution of dementia and cognitive impairment in PARDoS.

## Clinical-pathologic associations in 65+ decedents

### Single or mixed pathologies in decedents with dementia, MCI and NCI

We first focus on the clinical-pathologic associations among those age 65+ to compare our results to the literature. Supplementary figure 4 shows the distribution of neuropathologies alone or in combination in age 65+ decedents with dementia, MCI and NCI using the diagnostic pathologic thresholds of moderate to severe disease. More than 85% of persons with dementia had one or more of the four pathologies with more than half having mixed pathologies, most of which were AD plus one or more other pathologies. About three quarters of those with MCI had one or more brain pathologies, of whom about half had a single pathology and the remainder had mixed pathologies. In NCI, nearly 60% had one or more pathologies, more than half of which was a single pathology.

### AD and other neuropathologies in decedents with and without dementia

Intermediate to high ADNC, LBD and moderate to severe CAA were two times more frequent in dementia. Gross infarcts were found in more than two-fifths of dementia decedents, while in more than a quarter of decedents without dementia (all p<0.001). Table 2 shows the ADRD differences in 65+ decedents with and without dementia.

**Table 2.** Neuropathologic characteristics by dementia status in 65+ and 18-64 decedents.

| CHARACTERISTICS | 65+ decedents |  |  |  | 18-64 decedents |  |  |  |
| --- | --- | --- | --- | --- | --- | --- | --- | --- |
|  | N | With<br>Dementia | Without<br>Dementia | P | N | With<br>Dementia | Without<br>Dementia | P |
| Thal Staging $\geq 3$, n (%) | 3436 | 714 (61.2) | 783 (34.5) | <0.001 | 1471 | 13 (12.1) | 114 (8.4) | 0.18 |
| Braak Staging $\geq 3$, n (%) | 3438 | 1009 (86.3) | 1466 (64.6) | <0.001 | 1471 | 22 (20.9) | 171 (12.5) | 0.01 |
| Moderate to Severe Phosphorylated Plaques, n (%) | 3434 | 443 (37.9) | 219 (9.7) | <0.001 | 1451 | 4 (3.7) | 13 (1.0) | 0.01 |
| Intermediate to High ADNC, n (%) | 3436 | 715 (61.3) | 694 (30.6) | <0.001 | 1471 | 8 (7.5) | 44 (3.2) | 0.02 |
| Limbic and Neocortical LBD, n (%) | 3425 | 279 (23.9) | 252 (11.1) | <0.001 | 1454 | 9 (8.4) | 40 (3.0) | 0.003 |
| Gross Infarcts, n (%) | 3484 | 504 (42.7) | 623 (27.0) | <0.001 | 1482 | 37 (34.3) | 287 (20.9) | 0.001 |
| Moderate to Severe CAA, n (%) | 3465 | 467 (39.6) | 474 (20.7) | <0.001 | 1477 | 10 (9.3) | 69 (5.0) | 0.06 |

### Association of AD and other neuropathologies with dementia and cognitive impairment

The associations of the three AD indices used to derive ADNC with dementia are shown in Supplemental material. Intermediate to high ADNC, LBD, chronic gross infarcts and moderate to severe CAA were associated with dementia in separate models adjusted for demographics (all p<0.001) with effects sizes similar to 16 years of aging for intermediate to high ADNC, 12 years for LBD, 10 years for moderate to severe CAA and 9 years for chronic gross infarcts. All four neuropathologies remained associated with dementia when added to a single model adjusted for demographics with similar effect sizes (Figure 3 and Table 3) ranging from an odds ratio of 2.3 for pathologic AD and LBD to 1.4 for CAA. Decedents with all four pathologies had an OR for dementia of 7.2 (CI=3.4-15.1).

**Figure 3.**
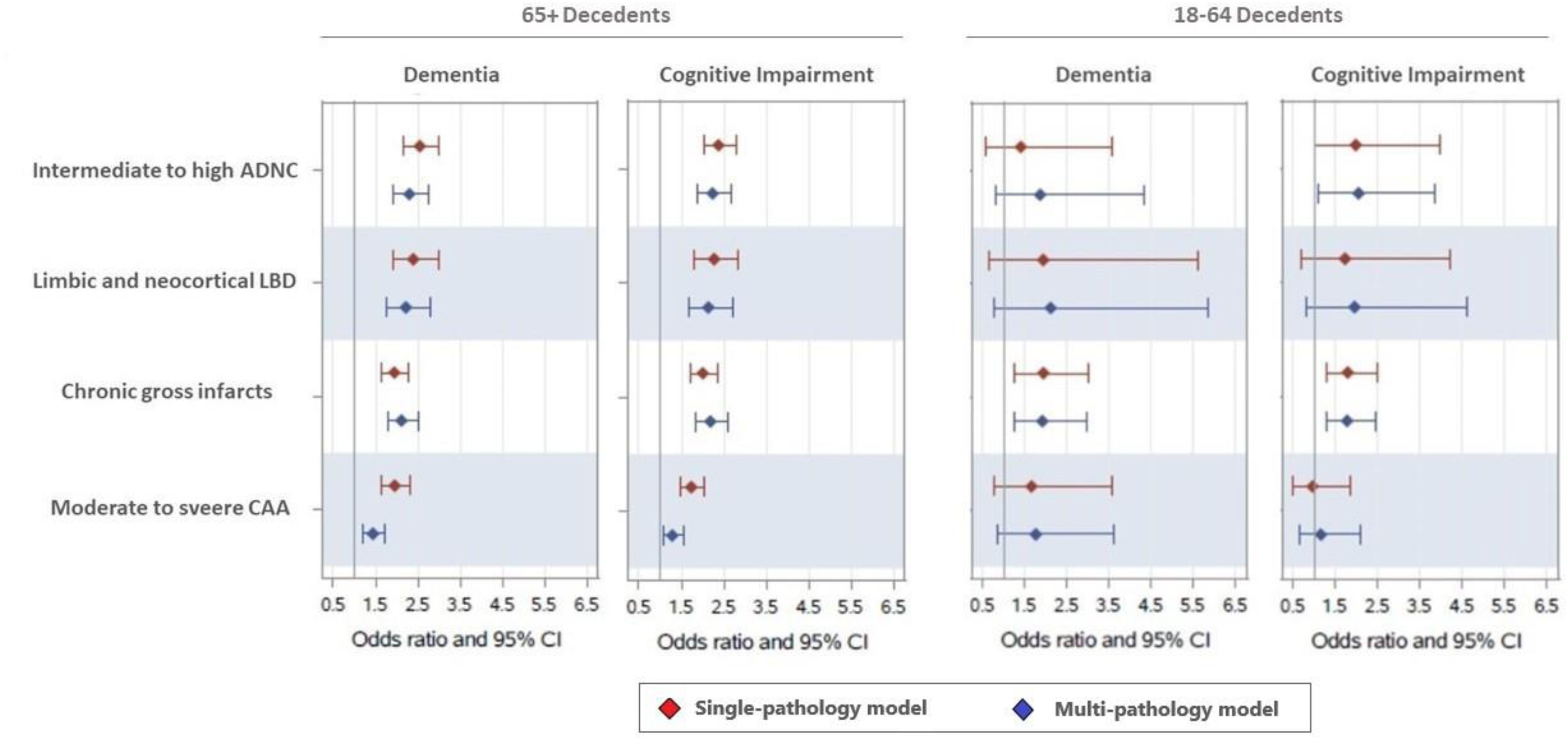
Associations of neuropathologies with dementia and cognitive impairment in 65+ and 18-64 decedents. Odds ratios are from logistic regression models adjusted for age at death, sex, race, and education. ‘Single-pathology models’ include each neuropathology individually. ‘Multi-pathology models’ include all the neuropathologies shown in the figure.

**Table 3.** Association of neuropathologies with dementia and cognitive impairment in 18-64 and 65+ decedents.

|  | 65+ Decedents (N=3362), OR (95%CI), p |  | 18-64 Decedents (N=1456), OR (95%CI), p |  |
| --- | --- | --- | --- | --- |
|  | Dementia | Cognitive Impairment | Dementia | Cognitive Impairment |
| Intermediate to High ADNC | 2.35 (1.97-2.80), p<0.001 | 2.28 (1.92-2.70), p<0.001 | 1.56 (0.63-3.88), p=0.34 | 2.19 (1.12-4.30), p=0.02 |
| Limbic and Neocortical LBD | 2.35 (1.88-2.94), p<0.001 | 2.18 (1.73-2.75), p<0.001 | 1.66 (0.59-4.69), p=0.33 | 2.01 (0.89-4.51), p=0.09 |
| Chronic Gross Infarcts | 2.16 (1.83-2.56), p<0.001 | 2.15 (1.83-2.52), p<0.001 | 1.88 (1.22-2.91), p=0.005 | 1.77 (1.29-2.44), p<0.001 |
| Moderate to Severe CAA | 1.42 (1.18-1.71), p<0.001 | 1.28 (1.06-1.53), p=0.008 | 1.28 (0.59-2.80), p=0.54 | 0.85 (0.45-1.61), 0.62 |

In separate logistic regression model with dementia presence or cognitive impairment presence as the outcome and controlling for age at death, sex, race and education with all four neuropathologies on table entered simultaneously ("multi-pathology model")

Supplemental material describes the associations of the three AD indices used to derive ADNC with cognitive impairment. Intermediate to high ADNC, chronic gross infarcts, LBD, moderate to severe CAA were also associated with cognitive impairment in separate models adjusted for demographics (all p<0.001) with effects sizes similar to 15 years of aging for ADNC, 10 years for chronic gross infarct and LBD and 8 years for CAA. All the four neuropathologies remained associated with cognitive impairment when added to a single model adjusted for demographics with similar effect sizes, except for CAA for which the effect size was reduced by half (Figure 3 and Table 3).

### Clinical-pathologic associations in the 18-64 decedents

The associations in those age 65+ are similar to findings in other community-based clinical-pathologic studies. This lends high confidence in the clinical and pathologic processes implemented to accomplish a study of this magnitude in such a compressed timeline. Next, we turn to the nearly 1500 persons age 18-64 which to the best of our knowledge, has never been the subject of a community-based clinical-pathologic study.

### Single or mixed pathologies in decedents with dementia, MCI and NCI

In decedents age 18-64, a single pathology was more common than a combination of pathologies in dementia, MCI and NCI brains (Supplementary Figure 4). In dementia and MCI, a single pathology was found in nearly two-fifths and a third of the brains, respectively while mixed pathologies were present only in one-tenth of the brains. In NCI, a quarter of the brain had a single pathology while less than 5% had a combination of pathologies. Brain infarcts alone accounted for most of the cases with neuropathology in the 18-64 dementia, MCI and NCI decedents. More than half of the dementia, three-fifths of the MCI and three quarters of the NCI decedents had none of the four pathologies.

### AD and other neuropathologies in decedents with and without dementia

Intermediate to high ADNC was two times and LBD three times more frequent in dementia. Gross infarcts were found in a third of dementia decedents, while in one-fifth of decedents without dementia. There were no statistically significant differences for CAA. Table 2 shows ADRD frequencies in 18-64 decedents with and without dementia.

### Association of AD and other neuropathologies with dementia and cognitive impairment

The associations of the three AD indices used to derive ADNC with dementia are shown in supplementary material. Intermediate to high ADNC were not associated with dementia in 18-64 brains. Chronic gross infarcts, more than 4.5 times more common than intermediate to high ADNC, were the only neuropathology associated with dementia with an effect size similar to 10 years of aging both in models adjusted for demographics and models adjusted for demographics and other neuropathologies (Figure 3 and Table 3).

Supplemental material shows the associations of the three AD indices used to derive ADNC with cognitive impairment. Different than the results obtained for dementia, intermediate to high ADNC was modestly associated with cognitive impairment in addition to chronic gross infarcts with effect sizes similar to 20 and 15 years of aging, respectively (Figure 3 and Table 3).

## Discussion

In this community-based clinical-pathologic study of 5300+ diverse Brazilians aged 18-108 we showed the age-distribution of dementia, cognitive impairment, four AD neuropathologic indices, and three additional common neuropathologies across eight decades. We examined the frequency of four common ADRD alone and in combination in dementia, MCI and NCI and showed differences in ADRD frequencies in young and older decedents with and without dementia. Finally, we examined the association of AD and the other three common neuropathologies with dementia and cognitive impairment in brains of young and older adults.

Multiple pathologies, including the ones examined in this study, were previously associated with a higher odds of dementia and cognitive impairment in the very-old [4,17–27]. Consistent with these previous studies, we show that AD, LBD, chronic gross infarcts and CAA were highly associated with both dementia and cognitive impairment in older adults. Further, the three indices used to generate ADNC were associated with dementia and cognitive impairment in 65+ decedents lending confidence to our clinical and neuropathologic assessments. Several of the prior clinical-pathologic studies reported that mixed pathologies, most frequently involving AD, are the most common cause of dementia in older adults. [4–8] Our findings mirror the latter findings, with more than half of the brains of older decedents with dementia showing intermediate or high ADNC, most commonly in combination with one or more pathologies. As expected and similar to previous work, the frequency of mixed pathologies is lower among those with MCI and NCI with more brains showing a single pathology than a combination of them. [5]

With the exception of another large study also conducted in Brazil [26], community-based clinical-pathologic studies mostly enroll highly educated, non-Latino white women, very-old participants with autopsy being performed after the age of 85 and few brains being examined before the eighth decade. [4,13–20,22,23] Studies with biomarkers for AD, including brain imaging and fluid biomarkers, were conducted in young adults, but included relatively a small sample sizes and were mostly restricted to subjects with high genetic risk for AD. [36–38] To the best of our knowledge, this is the first community-based clinical-pathologic study reporting on the relation of common aging brain pathologies to dementia and cognitive impairment among persons age 18-64. Here, we show that the two most frequent neuropathologies in our study, AD and chronic gross infarcts, are seen much earlier than when most of the clinical-pathologic studies start enrollment. β-amyloid deposits sufficient for Thal stage ≥ 3, PHFtau tangles sufficient for Braak stage ≥ 3, and intermediate to high ADNC consistent with a pathologic AD are first observed in the fourth decade. Chronic gross infarcts are already present in the early third decade and identified in more than one-tenth of the brains in the fourth decade. These novel findings reveal how early degenerative and vascular neuropathologies related to dementia develop in the brain. Although dementia is more common among older adults, the pathologies underlying cognitive impairment start in young adults, similar to what previous autopsy studies have reported for age-related chronic diseases in other organs [39,40]. These findings reinforce the importance of community-based clinical-pathologic studies including diverse samples with a broader age-range and are relevant from a public health perspective providing evidence that to be effective, primary and secondary prevention of ADRD likely needs to start in mid-life.

Further, our study markedly extends prior literature by examining the pathologies underlying dementia, MCI and NCI in young adults. Our results were strikingly different than seen for those age 65+. Mixed pathologies are much less common and a single pathology, namely chronic gross infarcts, dominated those with dementia being more than 4.5 times more common than intermediate to high likelihood ADNC. Chronic gross infarcts were the only pathology associated with dementia in young adults and were strongly associated with cognitive impairment with a modest additional effect from ADNC. These novel data illustrate the impact of vascular disease on dementia in younger racially and socioeconomically diverse Brazilians. Further studies are needed to better characterize the age, location and size of the gross infarcts and to add other vascular phenotypes collected in PARDoS such as atherosclerosis, arteriolosclerosis and microinfarcts to fully understand the role of vascular cognitive impairment in younger adults.

This study has multiple strengths. The participants are recruited in the community avoiding biases related to referred cases with unusual presentations at tertiary care clinics. PARDoS includes a diverse sample regarding age, sex, race, education and socioeconomic status increasing the ability to generalize the results to a larger population. The unique age distribution ranging from 18 to 108 years with about 1500 under age 65 is very unique and allows us to understand when these neuropathologies begin to accumulate in brain and see that clinical-pathologic relationships differ for those under age 65 relative to older persons. Finally, we were able to analyze very large numbers of brains relative to other clinical-pathologic studies. This study also has limitations. PARDoS is an autopsy-based study so the younger persons are not typical of living young persons and are probably enriched with some diseases of interest, in particular infarcts. It also does not allow for conclusions on the progression of neuropathologies over time as the subjects who died at an earlier age are not comparable to those who died later in life. Thus, the frequencies of conditions and diagnoses cannot be compared to frequencies reported in epidemiological studies. Participants were not tested during life and we rely on knowledgeable informants. The informant-based approach to clinical diagnoses is standard in the field and shows good correlation with antemortem in person diagnoses [41–43]. Other common neuropathologies including LATE-NC, hippocampal sclerosis, microinfarcts, arteriolosclerosis and others have also been associated with dementia and were not examined in the current study and will be reported separately. Chronic gross infarcts were not confirmed by microscopy in this study, although we only included chronic infarcts diagnosed as definite on gross examination. The microscopic confirmation is also ongoing and will be reported at a later date.

## Data Availability

The data generated in the present study can be requested at www.radc.rush.edu

## Acknowledgement

The authors thank the thousands of Brazilian representatives who participated in this study and PARDoS staff in Brazil and USA. We thank the Rush Alzheimer’s Disease Center, the Núcleo de Estudos, Pesquisa e Assessoria à Saúde (NEPAS), the Autopsy Services (SVO) at Santo André and Guarulhos, and the Instituto de Assistência Médica ao Servidor Público Estadual (IAMSPE).

PARDoS is supported by NIA grants R01AG54058 and R01AG075927.

PARDoS resources can be requested at www.radc.rush.edu. The data repository is at the RADC and requires a Data Use Agreement (DUA) with RUMC which includes language making it compliant with Brazilian law. The PARDoS biorepository is in IAMSPE. Material can only be sent abroad or used with money from a foreign country with approvals from a Brazilian local ethical committee (CEP) and the national ethical committee (CONEP). Such approvals can only be granted to a Brazilian investigator affiliated with an institution in Brazil. Thus, access to PARDoS resources can only be done in collaboration with a Brazilian investigator and institution. A Material Transfer Agreement (MTA) will be required with IAMSPE. Both the DUA and the MTA are tailored to Brazilian laws and regulations and cannot be negotiated.

The investigators have no relevant conflicts of interest.

---

**Supplementary Table 1.** Association of AD indices with dementia and cognitive impairment in 65+ decedents.

|  | 65+ Decedents (N=3360), OR (95%CI), p, |  |  |  |
| --- | --- | --- | --- | --- |
|  | Dementia |  | Cognitive Impairment |  |
|  | Single-pathology models | Multi-pathology model | Single-pathology models | Multi-pathology model |
| Thal stage $\geq 3$ | 2.29 (1.99-2.68), p<0.001 | 1.37 (1.14-1.64), p<0.001 | 2.09 (1.80-2.42), p<0.001 | 1.37 (1.15-1.62), p<0.001 |
| Braak stage $\geq 3$ | 2.07 (1.69-2.54), p<0.001 | 1.45 (1.16-1.80), p<0.001 | 1.67 (1.39-1.99), p<0.001 | 1.22 (1.01-1.47), p=0.04 |
| Moderate to Severe Phosphorylated Plaques | 4.58 (3.78-5.56), p<0.001 | 3.54 (2.84-4.41), p<0.001 | 4.04 (3.29-4.96), p<0.001 | 3.24 (2.58-4.08), p<0.001 |

In separate logistic regression model with dementia presence or cognitive impairment presence as the outcome and controlling for age at death, sex, race and education with each neuropathology on table entered separate ("single-pathology model") and simultaneously ("multi-pathology model")

**Supplementary Table 2.** Association of AD indices with dementia and cognitive impairment in 18-64 decedents.

|  | 18-64 Decedents (N=1456), OR (95%CI), p, |  |  |  |
| --- | --- | --- | --- | --- |
|  | Dementia |  | Cognitive Impairment |  |
|  | Single-pathology models | Multi-pathology model | Single-pathology models | Multi-pathology model |
| Thal stage $\geq 3$ | 1.11 (0.58-2.11), p=0.75 | 0.96 (0.47-1.95), p=0.91 | 1.83 (1.19-2.81), p=0.006 | 1.67 (1.05-2.65), p=0.03 |
| Braak stage $\geq 3$ | 1.34 (0.79-2.27), p=0.28 | 1.23 (0.70-2.15), p=0.47 | 1.39 (0.94-2.04), p=0.10 | 1.17 (0.78-1.76), p=0.46 |
| Moderate to Severe Phosphorylated Plaques | 2.60 (0.72-9.46), p=0.15 | 2.37 (0.56-9.99), p=0.24 | 2.79 (0.99-7.84), p=0.06 | 1.70 (0.55-5.21), p=0.36 |

In separate logistic regression model with dementia presence or cognitive impairment presence as the outcome and controlling for age at death, sex, race and education with each neuropathology on table entered separate ("single-pathology model") and simultaneously ("multi-pathology model")

**Supplementary Figure 1.**
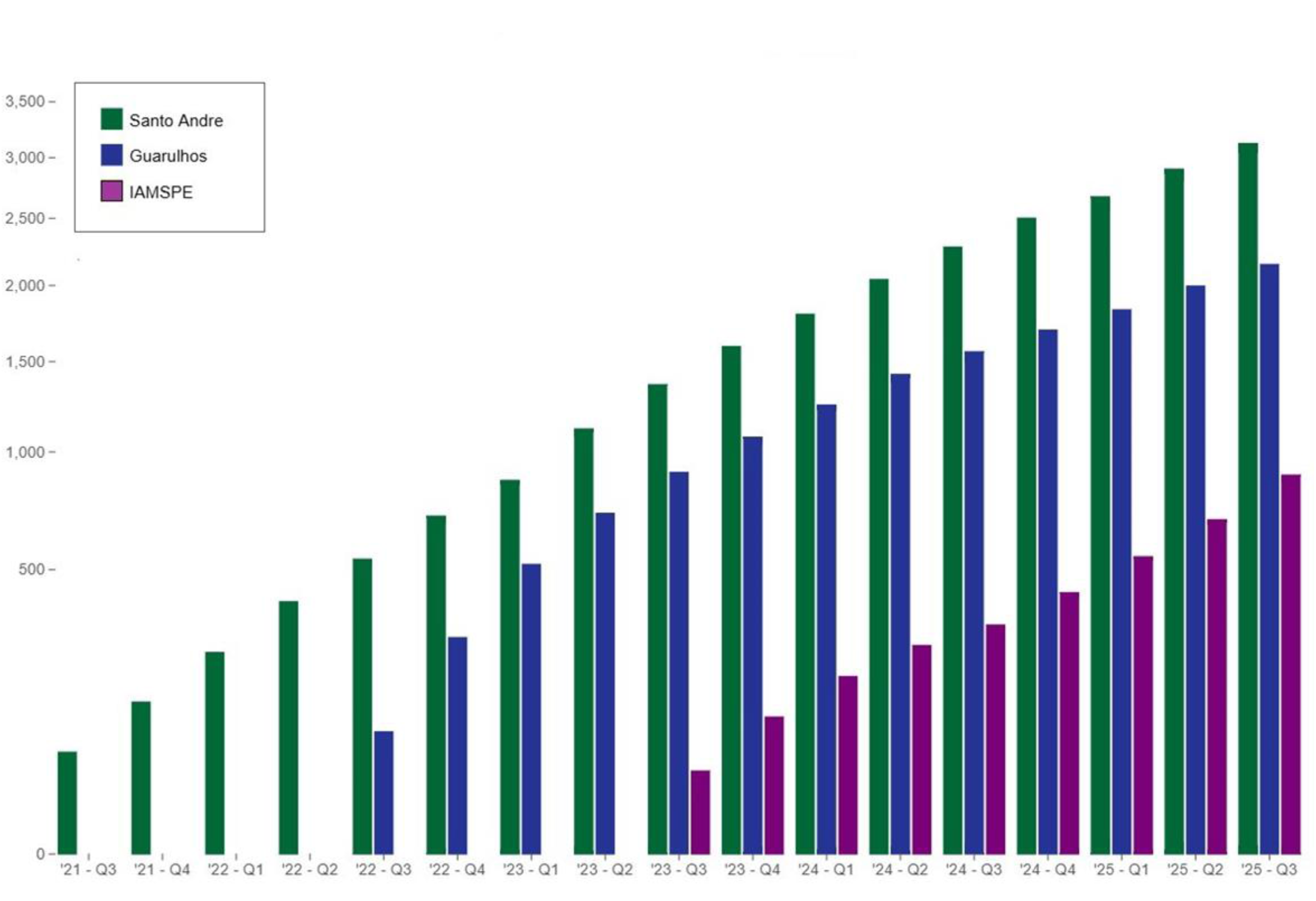
Brains collected in PARDoS three sites from July 2021 to September 2025.

**Supplementary Figure 2.**
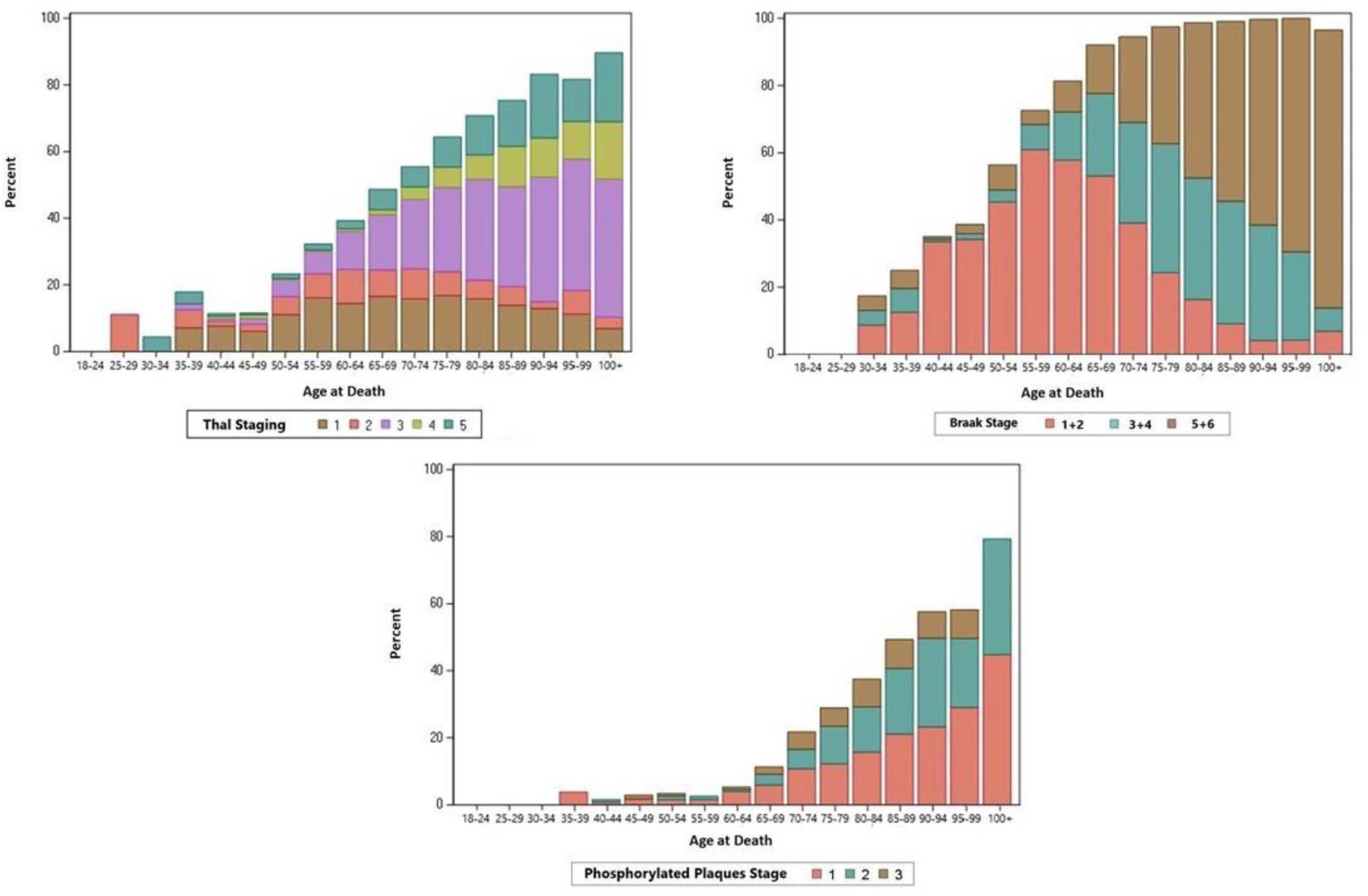
Age-distribution of three Alzheimer’s disease pathologic indices.

**Supplementary Figure 3.**
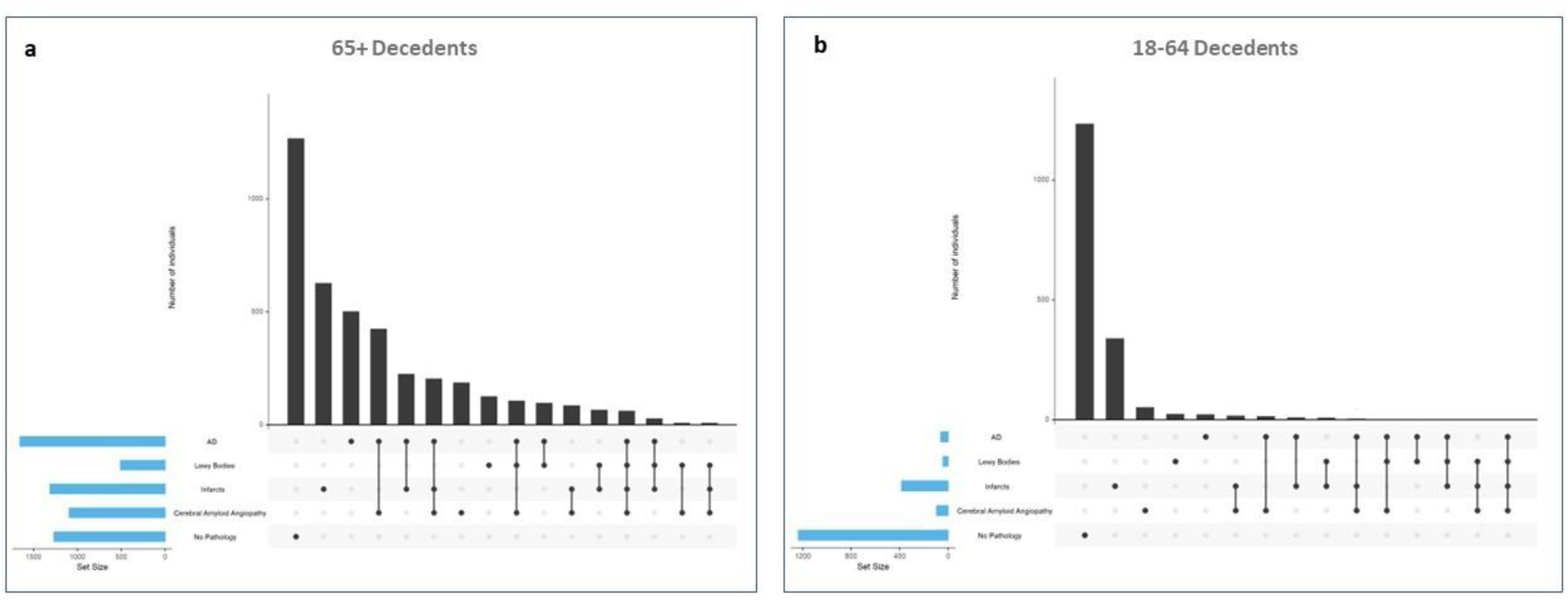
Distribution of neuropathologies alone and in combination in 65+ and 18-64 decedents.

**Supplementary Figure 4.**
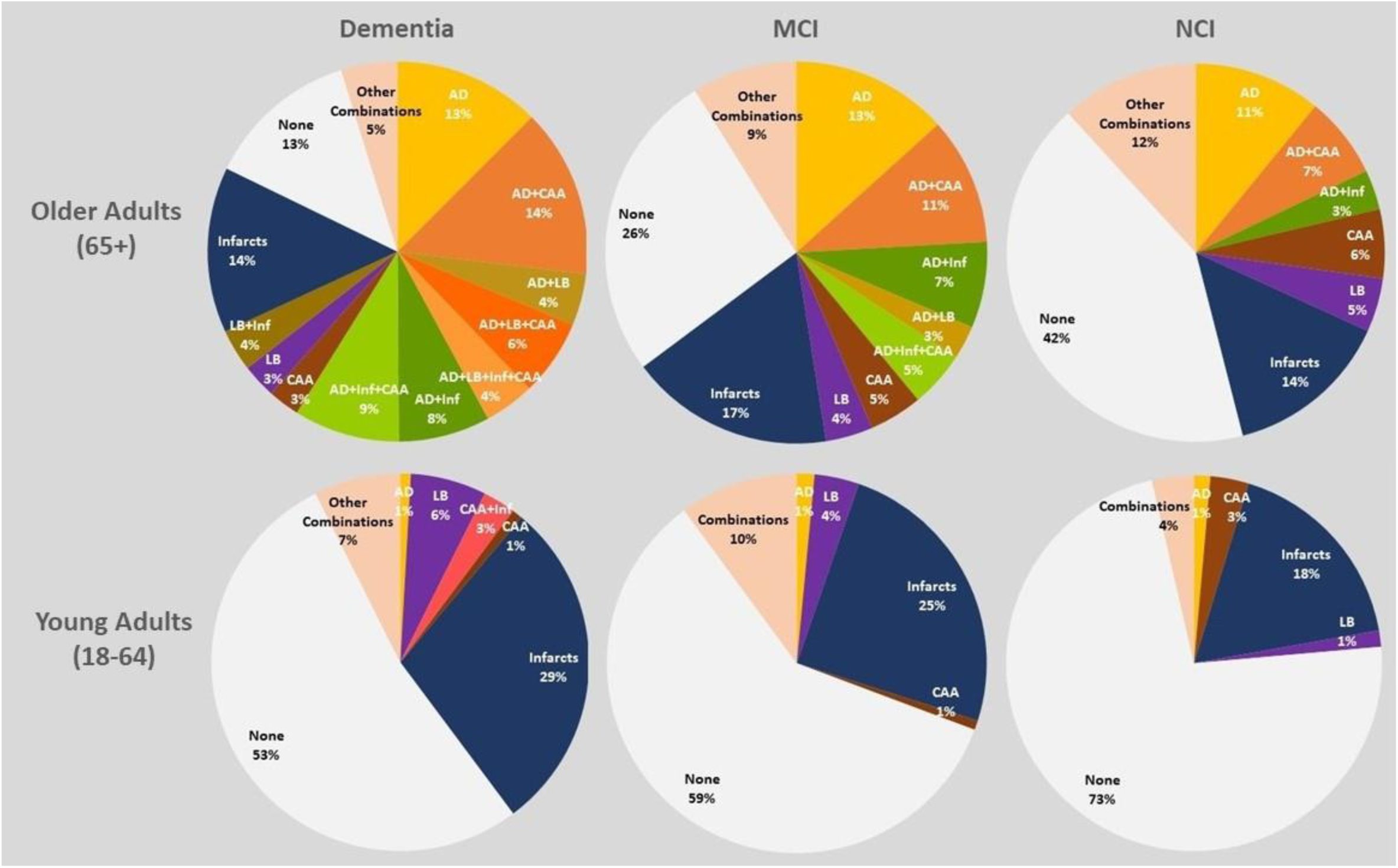
Distribution of neuropathologies alone and in combination in dementia, MCI and NCI.

## Supplemental Material for the manuscript “Age and the relation of common neuropathologies to dementia in Brazilian adults”

### Age-distribution of three AD pathological indices

β-amyloid deposits sufficient to contribute to a pathological diagnosis of AD (Thal stage ≥3) were first seen in the fourth decade, were present in nearly 5% in the fourth and fifth decades, one-tenth in the sixth, one-fifth in the seventh, two-fifths in the eighth decade and more than half of the brains afterwards. Neurofibrillary tangles in density and distribution sufficient to contribute to pathological AD (Braak stage ≥3) also were first seen in the fourth decade. Interestingly, they were present in up to 10% of the brains from the fourth decade to the sixth decade, then increased rapidly to one-fifth in the seventh, three-fifths in the eighth and over 90% in the ninth and tenth decades. Moderate to severe neocortical PPs were first found in the fifth decade, a decade following Thal > 2 and Braak > 2. They were rare before the age of 65 following which they increased rapidly with age to 5% of the 65-69 decedents, 20% in the eighth decade and a quarter afterwards. The age-distributions of the three AD indices used to derive ADNC can be found in Supplementary Figure 2.

Age-distribution of initial signs of three AD pathological indices

Early signs of AD pathology in the form of mild β-amyloid deposits (Thal stage 1-2) were found in the mid-third decade, present in more than one-tenth in the mid-fourth and fifth decade, a quarter between the sixth and eighth decades and decreased thereafter due to the large numbers with Thal > 2. Braak 1-2 was first seen in the first half of the fourth decade. It was present in 10% of brains in the fourth, a third in the fifth, half in the sixth and seventh decades and decreased thereafter due to the large numbers with Braak>2. Sparse PPs were first found in the second half of the fourth decade, were rare before the seventh decade, remained present in less than 10% in the seventh, one-tenth in the eighth, one-fifth in the ninth, a quarter in the tenth and two-fifths after the age of 100 (Supplementary Figure2). Low ADNC was seen as early as the late third decade, found in one-tenth of the brains from the mid-third to the fifth decade, a quarter in the sixth, a third in the seventh and decreased thereafter due to greater stages of ADNC (Figure 1a).

Olfactory LBs alone were first seen in the sixth decade and remained rare across the age spectrum. Nigra predominant LBD was initially seen in the early 40s and increased slightly leveling off at about 5% from the fifth to the tenth decade. Mild CAA was first seen in the 25-30 age group, in about 10% of the brains from the mid-third to the seventh decade and one-fifth afterwards (Figure 1b-d).

### AD indices in 65+ decedents with and without dementia

Table 2 shows the differences in frequency of the three AD indices in 65+ decedents with and without dementia. Thal stage ≥ 3 was two-times, Braak stage ≥3 was 20% greater and moderate to frequent neocortical PPs were four times more frequent in dementia decedents (all p<0.001).

### Association of the three AD indices AD with dementia in 65+ decedents

Logistic regression models examined the association of each of the three AD indices used to define ADNC with dementia in 65+ decedents, adjusted for demographics (Supplementary table 1). All the three AD indices were associated with dementia (all p<0.001) with effect sizes equivalent to 13 years of aging for Thal ≥3, 11 years for Braak ≥ 3, and 25 years for moderate to severe PPs. When added to a single model, all three pathologies remained statistically associated with dementia, but Thal ≥3 and Braak ≥ 3 had attenuated effects equivalent to 5 years of aging while moderate to severe PPs was effectively unchanged.

### Association of three AD indices with cognitive impairment in 65+ decedents

All three AD indices used to define ADNC were also associated with cognitive impairment in 65+ decedents when entered in separate models adjusted for demographics (all p<0.001) with odds equivalent to 11 years of age for Thal ≥3, 8 years for Braak ≥3 and 22 years for PPs. When the three indices were included in the same model, Braak stage ≥3 remained marginally associated with cognitive impairment with effect sizes similar to 3 years of aging, Thal stage ≥3 had effects attenuated by half, and PPs remained associated with cognitive impairment essentially unchanged (Supplementary Table 1).

### AD indices in 18-64 decedents with and without dementia

Among 18-64 decedents, Braak stage ≥3 was nearly two times and moderate to frequent PPs nearly four times more frequent in dementia decedents, respectively. We did not find statistically significant differences for Thal stage ≥ 3 (Table 2)

### Association of the three AD indices AD with dementia in 18-64 decedents

None of the three AD indices were associated with dementia in 18-64 decedents (Supplementary table 2)

### Association of three AD indices with cognitive impairment in 18-64 decedents

Supplementary table 2 describe the associations of the three AD indices used to derive ADNC with cognitive impairment. Among the three indices used to define ADNC, Thal ≥3 was associated with cognitive impairment with effects similar to 15 years of aging in separate models adjusted for demographics and models adding the two other AD indices. We didn’t find an association of Braak ≥ 3 and moderate to severe PPs with cognitive impairment in 18-64 decedents.

